# Immunologically Optimized Zmp1 Peptides Reveal a Translational Serological Biomarker Platform for Tuberculosis Diagnosis Across Disease Manifestations

**DOI:** 10.64898/2026.06.11.26355355

**Authors:** Omkar Shriram Zade, Sriram Yandrapally, Keerthana Choudari, Akshay Vaijinath Gaikwad, Rajanikant Panda, Venkata Sanjeev Kumar Neela, Devalraju Kamakshi Prudhula, V. V. Ramprasad Eedara, Mohammad Soheb Ansari, Chitra Chandrashekhar, Divya Sriram, Krishnaveni Mohareer, Vijaya Lakshmi Valluri, Pramod Rajaram Somvanshi, Sharmistha Banerjee

**Author notes:** Yale University, New Haven, CT, USA-06511. CSIR-Centre for Cellular and Molecular Biology (CSIR-CCMB), Hyderabad, Telangana, India-500007. Contributed equally (Akshay Vaijinath Gaikwad, Rajanikant Panda). Contributed Equally (Venkata Sanjeev Kumar Neela, Devalraju Kamakshi Prudhula, V. V. Ramprasad Eedara).

## Abstract

Tuberculosis (TB) diagnosis remains challenging, particularly for extrapulmonary TB (EPTB), where invasive sampling, low bacillary burden, and suboptimal sensitivity of nucleic acid-based tests in peripheral specimens hinder timely detection. Here, we report an immunology-driven strategy for biomarker discovery and development of a peptide-based serological assay targeting *Mycobacterium tuberculosis* zinc metalloprotease-1 (Zmp1). Leveraging fundamental principles of adaptive immunity that antigenic regions containing overlapping B-cell and CD4□ T-helper cell epitopes would preferentially generate high antibody titers through linked recognition and cognate T-cell help, we used an immunoinformatics pipeline to identify two nested immunodominant peptide regions within Zmp1 (Mtb-Zp-NT and Mtb-Zp-CT) enriched for overlapping B- and T-cell epitopes.

The diagnostic potential of these peptides was evaluated through ELISA-based serological assays. A blinded pilot study (N=137) demonstrated a clear discrimination between active TB and TB-recovered individuals. The assay was subsequently validated in an expanded cohort (N=875) by screening 6,086 individuals, which identified 457 TB-positive cases. The cohort included pulmonary TB (PTB), EPTB, TB-recovered individuals, household contacts, non-specific infections, and healthy controls. Receiver operating characteristic analyses, supported by DeLong and bootstrap comparisons, revealed superior diagnostic performance of the peptide-based assays relative to full-length Zmp1. Mtb-Zp-CT exhibited the highest accuracy (AUC=0.93; specificity >90%), while Mtb-Zp-NT also demonstrated strong discriminatory power (AUC≈0.89).

These findings establish that the immunologically optimized Zmp1 peptides are highly promising serological biomarkers for TB and EPTB. More broadly, they demonstrate how mechanistically informed epitope selection can accelerate translation of pathogen-specific immune signatures into sensitive, minimally invasive, and potentially point-of-care diagnostic platforms for resource-limited settings.

**Key points:**

- We identified two peptides from M.tb Zmp-1 protein that elicit strong, active TB□specific antibody responses, including hard□to□diagnose extrapulmonary TB, to be detected using a simple blood test.
- In both the pilot study as well as large multi-centre cohort, these peptides showed high accuracy and clearly distinguished active TB patients from healthy individuals and other infections, making them promising candidates for future point□of□care tests.

## 1. Introduction

Tuberculosis (TB), caused by *Mycobacterium tuberculosis* (*M*.*tb*), remains a leading infectious disease worldwide, with India bearing the highest burden. Beyond pulmonary disease, TB can present as extrapulmonary TB (EPTB), which is often difficult to diagnose and disproportionately affects children, women, older adults, and immunocompromised individuals. Ongoing transmission, driven by poverty, undernutrition, and overcrowding, together with the rise of drug-resistant *M*.*tb* strains, underscores the need for rapid, accurate, and accessible diagnostic tools across the spectrum of TB [1].

The clinical heterogeneity of TB, reflected by variations in bacterial burden, tissue involvement, and host immune responses across PTB and EPTB, limits the performance of current diagnostic tools. Serological and antigen-based assays show poor sensitivity in paucibacillary and extrapulmonary disease [2–4], resulting in missed diagnoses among patients with EPTB, smear-negative TB, and HIV-associated TB [5]. Diagnosis is further hindered by nonspecific clinical manifestations, low bacillary loads, and the frequent need for invasive sampling from extrapulmonary sites [6,7]. Although nucleic acid amplification tests such as GeneXpert MTB/RIF have improved TB detection, their dependence on invasive specimens for EPTB and laboratory infrastructure restricts use in many high-burden settings in remote areas [8]. Their sensitivity is also limited in peripheral samples such as blood and urine, where *M*.*tb* DNA is often absent or scarce [9]. Other methods suffer from important drawbacks, including low sensitivity of smear microscopy, nonspecific radiographic findings, prolonged culture turnaround times, and confounding of the tuberculin skin test by BCG vaccination. Collectively, these limitations highlight the need for rapid, affordable, non-sputum-based diagnostics capable of detecting both PTB and EPTB from readily accessible biospecimens such as oral swabs, urine, and serum.

Serology offers a minimally invasive diagnostic strategy for TB, particularly EPTB. Earlier assays based on whole-cell lysates or full-length antigens lacked specificity owing to epitope masking, antigen heterogeneity, and cross-reactivity with non-tuberculous mycobacteria, leading to the WHO’s non-endorsement of commercial serological tests [10]. Nonetheless, a rapid point-of-care assay targeting a secreted, immunodominant antigen with disease-specific antibody responses could enable frontline screening and complement molecular confirmation.

We previously demonstrated that active TB elicits high-titre antibodies against the secreted virulence factor Zmp1, independent of bacillary burden or disease site, identifying anti-Zmp1 antibodies as a potential biomarker for both paucibacillary and extrapulmonary disease [11]. Leveraging the principle that antigenic regions containing overlapping B-cell and CD4□ T-cell epitopes are highly immunogenic [12], we used immunoinformatics to identify immunodominant nested epitopes within Zmp1 predicted to induce robust antibody responses. The diagnostic performance of the resulting peptides was first evaluated in a pilot cohort (n=137) and subsequently validated in an independent cohort (n=875) comprising PTB, EPTB, healthy controls (HC), household contacts (HHC), non-specific infections (NSI), and TB-recovered patients (TBR), and selected from 6,086 screened participants. ROC, DeLong, and bootstrap analyses demonstrated that these optimized Zmp1 peptides are promising serological biomarkers, particularly for EPTB, and support the development of a sensitive, minimally invasive point-of-care TB diagnostic.

## 2. Methods

### 2.1 Study design

The study design is schematically summarised in Figure 1. The study was divided into two stages. Stage 1 was a smaller cohort with a blinded sample set comprising active TB (including both PTB and EPTB), (HC), and (HHC). We included patients who had recovered from TB in stage 1 to rule out false positives and to assess circulating antibodies post-treatment at 0, 3, 6, 12, and 24 months. The observation of discriminatory antibody titers in positive samples compared to controls prompted us to advance to stage 2 of our study. In the expanded stage 2, we continued the work from stage 1, incorporating an additional control group with nonspecific infections to reduce diagnostic complexity.

**Fig 1.**
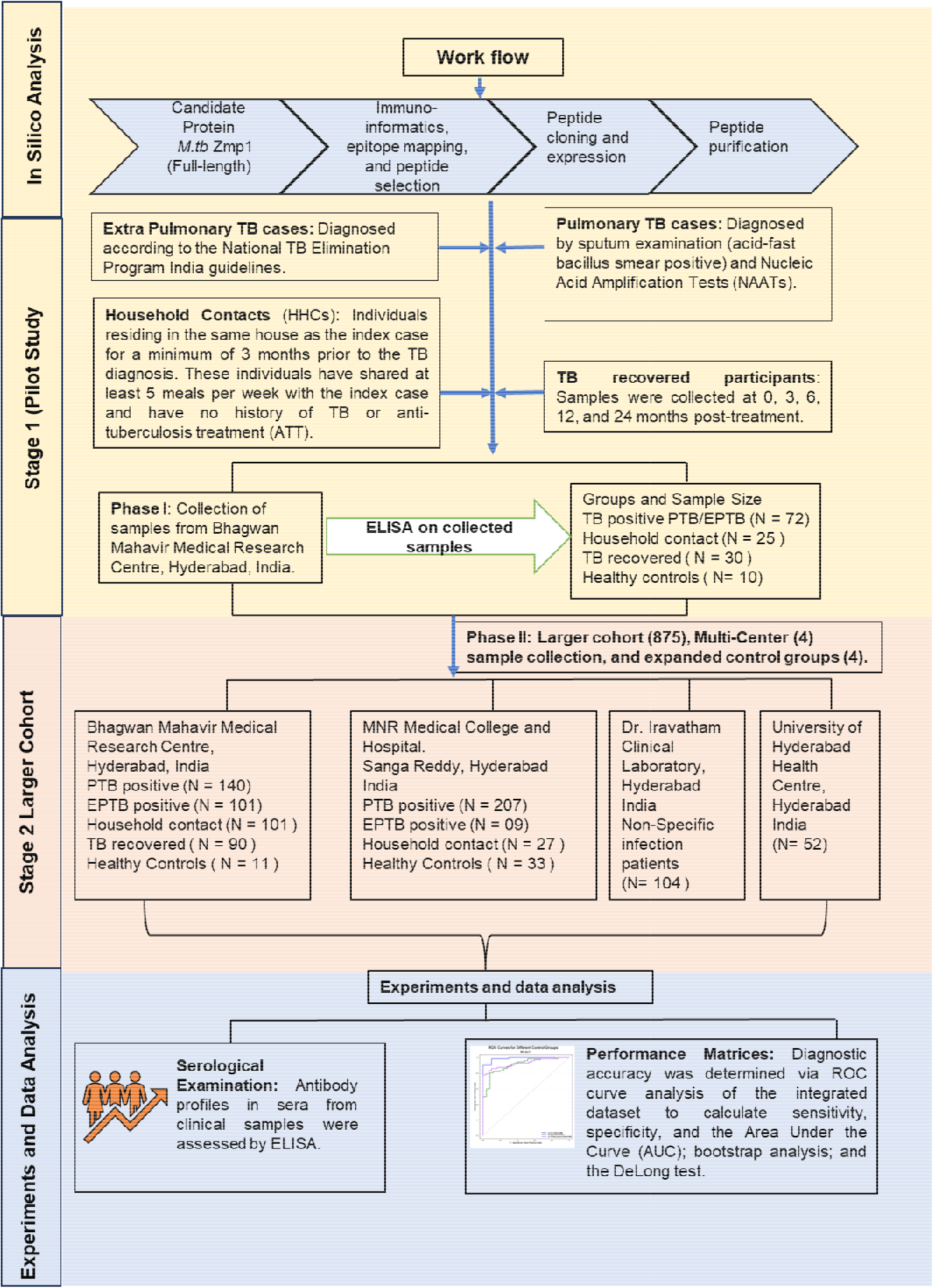
Schematic overview of the study design. Rationally designed peptides derived from the full□length Mtb□Zmp1 protein were generated through immunoinformatics□based epitope mapping, followed by peptide cloning, expression, and purification. The study was conducted in two Stages. Stage-1 comprised a pilot evaluation of the diagnostic performance of the designed peptides in a small cohort (N□=□137). Based on the promising results, Stage 2 was undertaken in a larger cohort recruited from multiple screening centers (N□=□875). Refer to the main text for detailed methodology. **Abbreviations in Fig 1** ***M*.*tb***: Mycobacterium *tuberculosis;* **Zmp-1**: Zinc Metallo Protease-1; **TB**: Tuberculosis; **N**: Sample number; **NAAT**: Nucleic Acid Amplification Test; **ELISA**: Enzyme-Linked Immuno Sorbent Assay; **HHC**: House Hold Contacts; **ATT**: Anti-Tuberculosis Therapy; **PTB**: Pulmonary Tuberculosis; **EPTB**: Extra-Pulmonary Tuberculosis; **ROC**: Receiver Operating Characteristics; **AUC**: Area Under Curve

### 2.2 Study cohort

#### 2.2.1 Stage-1

N=137, including active TB (N=72), HC (N=10), HHC (N=25), and TB-recovered individuals (N=30).

#### 2.2.2 Stage-2

The cohort was expanded to reflect real-world diagnostic complexity by separating active TB into PTB and EPTB and by adding NSI as an additional control. The sample size was calculated based on an expected AUC of 0.8, 90% power, and a significance level of α = 0.05 for a multicentric study design (supplementary methods: Multicentre Clustering Adjustment; Supplementary Table S1). Sera were obtained after screening 6086 (male-female ratio of 55:45) individuals with suspected TB at four clinical centres, yielding a study cohort of 875 participants: 457 active TB cases and 418 controls. Participants were classified into six groups: PTB (N=347), EPTB (N=110), HC (N=96), TBR (N=90), NSI (N=104), and HHC (N=128).

PTB diagnosis followed NTEP guidelines, incorporating sputum AFB microscopy, PCR-based detection, culture, chest radiography, and tuberculin skin testing [13]. EPTB was defined by culture positivity from extra-pulmonary sites or by histological, radiological, or strong clinical evidence of active disease. Household contacts were individuals residing with a TB patient for ≥7 consecutive days within the preceding three months [14]. Although asymptomatic, many were Mantoux positive, reflecting latent infection risk [13]. Non-specific infection controls included individuals with confirmed viral, bacterial, or parasitic infections other than TB [15]. Eligible TB patients were aged 10-75 years with symptoms consistent with TB and microbiological or radiological evidence of disease [14]. Exclusion criteria were pregnancy, age <8 or >75, and concurrent conditions or medications likely to alter immune responses.

### 2.3 Immuno-informatics for peptide prediction

#### 2.3.1 B-cell Epitope Prediction

Linear B-cell epitopes were identified using BepiPred-2.0 [16], which predicts epitope-rich regions from antigen sequences using a random forest model trained on antibody-antigen crystal structures. It outperforms earlier sequence-based tools using structural datasets and IEDB-curated linear epitopes. Predicted residues were visualized in PyMOL (Schrödinger, LLC) to interpret B- and T-cell epitope locations. Conformational B-cell epitopes were predicted with DiscoTope 2.0 [17], which identifies discontinuous epitopes in 3D structures by integrating residue surface accessibility, amino acid-specific propensity scores, and contact number-based spatial weighting. Predicted antigenic residues were mapped onto the protein structure.

#### 2.3.2 T-cell Epitope Prediction

MHC class-II T-cell epitopes were predicted using the IEDB pipeline. The Zmp1 sequence was screened across multiple HLA-II alleles; consistent epitopic regions allowed retention of two representative alleles. ∼650 candidate peptides were identified, including 37 strong MHC class-II binders and ∼600 weaker binders. Epitopes were visualised on the sequence and 3D structure. IEDB proteolytic cleavage/elution analysis identified five peptides predicted to bind MHC class-II and be generated during antigen processing in the MHC class-II presentation pathway.

### 2.4 Gene Cloning, Protein Expression and Purification

Rv0198c (Zmp1) gene and its fragments encoding the predicted N-terminal (Mtb-Zp-NT) and C-terminal (Mtb-Zp-CT) peptides were amplified from *M*.*tb* H37Rv genome, cloned and expressed in *E. coli* BL21 (DE3) cells. Recombinant His-tagged full-length Zmp1, Mtb-Zp-CT and Mtb-Zp-NT were purified under native and denaturing conditions (Mtb-Zp-NT), using Ni-NTA affinity chromatography (Qiagen). Dialysis was performed in 50 mM Tris-HCl (pH 8), 200 mM NaCl, and 5% glycerol for full-length Zmp1 and Mtb-Zp-CT. Mtb-Zp-NT underwent stepwise dialysis with decreasing urea concentrations for renaturation. SDS-PAGE analysis confirmed molecular weights of ∼75 kDa (Zmp1), 22 kDa (Mtb-Zp-CT), and 20 kDa (Mtb-Zp-NT).

### 2.5 Enzyme-Linked Immunosorbent Assay (ELISA)

ELISA was performed using standard immunoassay protocols. Microplates were coated with 100 ng protein/well in 100 µL carbonate buffer and incubated overnight at 4°C. Plates were washed thrice with 100 µL PBST, blocked with 10% FBS (in 1× PBS) for one hour at room temperature, and washed again thrice. Patient sera (1:200 in PBST) were added in triplicate, and the sealed plate was incubated for two hours at room temperature, followed by four PBST washes and incubation with HRP-conjugated anti-human secondary antibody (Invitrogen) for one hour. After seven washes with PBST, 50μl of TMB and 50μl of peroxidase buffer mixed to a total 100μl was added and incubated in the dark for 5-30 min until a blue colour was observed. Reactions were stopped with 2 N H□SO□, and absorbance was measured at 450 nm. This workflow aligns with established TB serology assay formats [18,19].

### 2.6 Statistical Analysis

All statistical analyses were performed using R software (version 2023.06.2, Mountain Hydrangea) in the RStudio environment [20]. We used three R packages mainly for data processing and analysis. The ‘readxl’ package enabled direct import of optical density values from Excel spreadsheets. The ‘dplyr’ package was used for data processing and organisation of samples into analytical groups. The ‘pROC’ package was used for receiver operating characteristic curve analysis, area under the curve calculation and threshold determination [21]. The Optical density values obtained from ELISA were grouped into five categories: active TB (PTB and EPTB), HC, HHC, NSI, and TBR. The Wilcoxon rank-sum test was used to compare optical density values between the groups [22]. Each active TB group (PTB and EPTB) was tested against all four control populations (HC, HHC, NSI, and TBR), and results were considered significant when p-values were below 0.05.

Receiver operating characteristic (ROC) curve analysis [23] was used to assess how well each antigen could separate TB cases from controls. The curve shows sensitivity (True Positive Rate) on one axis and false positive rate (1-specificity) on the other, letting us see how changes in the threshold affect both measures at once. We used the area under the curve to summarise overall performance. A value of 0.5 means the test performs no better than guessing, while 1.0 means perfect separation. The values above 0.7 are generally acceptable, above 0.8 indicates good performance, and above 0.9 suggests excellent diagnostic ability.

To find the best threshold for distinguishing positive from negative results, we used the Youden index. For each possible cut-off value along the ROC curve, the index is calculated as J = sensitivity + specificity − 1, where J denotes the Youden index. The calculation involves determining sensitivity and specificity at every threshold, then identifying the point where their sum minus one reaches its maximum. The optical density value corresponding to this maximum J value is selected as the optimal diagnostic cut-off. Samples with readings above this cut-off are classified as positive, while those below are classified as negative. For each comparison between the positive group and control populations, we recorded this optimal threshold along with the corresponding sensitivity and specificity values. Results were displayed using paired figures showing violin plots with box plots and significance markers on one side, and receiver operating characteristic curves with marked optimal points and area under the curve values on the other.

To assess the reliability and generalizability of our diagnostic thresholds, we performed bootstrap validation using 1000 iterations [24]. For each iteration, the dataset was randomly split into training (80%) and test (20%) sets. The optimal cut-off was determined using the Youden index on the training set, then applied to the independent test set to evaluate performance. This procedure was repeated 1000 times with a fixed random seed (123) to ensure reproducibility. We calculated mean values and 95% confidence intervals for area under the curve, sensitivity, specificity and accuracy across all iterations. Model performance was classified as robust when test AUC remained above 0.7 with confidence intervals excluding 0.5, acceptable when test AUC was above 0.65, or requiring review when below 0.65 or showing substantial performance drops between training and test sets. This validation strategy was applied separately to PTB and EPTB groups, each compared independently with each control group and also the overall TB-negative cohort, and was then extended to a combined analysis including all TB-positive cases versus all negatives to account for overall diagnostic heterogeneity.

## 3 Results

### 3.1 Conceptual framework of identifying B-cell and T-cell linked epitopes nested in *M*.*tb* antigen Zmp1 using integrative Immunoinformatics-Based Prediction

Given evidence of strong Zmp1-specific antibody responses in active TB, we performed in silico epitope mapping to identify diagnostically relevant regions of Zmp1. We used integrative immunoinformatics to systematically predict antigenic peptides incorporating B-cell and T-cell epitopes, enabling coordinated humoral and cellular immune recognition. Protein sequences were screened for antigenicity, conservancy, surface accessibility, and immunogenicity, followed by prediction of B-cell epitopes and MHC class I/II-restricted T-cell epitopes using computational algorithms. We prioritized “nested” regions in which B-cell and CD4□ T-cell epitopes overlap, facilitating linked recognition, affinity maturation, and robust antibody generation. This approach enriches for immunodominant peptide candidates with diagnostic potential while reducing experimental screening burden through in silico prioritization.

We used BepiPred-2.0 to predict linear B-cell epitopes and DiscoTope-2.0 to refine conformational candidates. This analysis identified two immunogenic hotspots: an N-terminal region (residues 148-298; Mtb-Zp-NT) and a C-terminal region (residues 445-601; Mtb-Zp-CT). Both regions showed high epitope propensity, surface accessibility, and favourable immunogenic characteristics (Supplementary Figure S1-A).

To assess CD4□ T-cell presentation, we analysed the Zmp1 sequence using the IEDB MHC-II pipeline, employing the NetMHCIIpan EL tool. Our scan of multiple HLA-II alleles identified about 650 candidate peptides ranging from 12 to 25 amino acids, including 37 strong binders with percentile ranks ≤2%. By applying processing and elution-aware predictors from the IEDB, we refined our selection to five peptides predicted to be strong binders, likely derived from natural endosomal and protease cleavage processes.

Mapping all predicted B- and T-cell epitopes onto the linear sequence and three-dimensional structure of Zmp1 revealed distinct clustering within residues 148-298 and 445-601 (see Supplementary Figure S1). These targeted regions were chosen for recombinant expression and purification, then evaluated alongside the full-length protein in downstream serological assays.

### 3.2 Stage 1 assessment: A blinded pilot study pointed to the ability of the peptides to significantly distinguish active TB from TB-recovered and household contacts

Stage 1 studies evaluated whether antibodies against Zmp1 and its peptides persist after TB recovery, a key requirement for diagnostic specificity. In a blinded pilot ELISA study on 137 samples from a single centre, Bhagwan Mahavir Medical Research Center, including active TB patients (n=72), HHC (n=25), TBR (n=30), and HC (n=10), were analyzed to assess baseline and disease-associated seroreactivity. Recombinant full-length Mtb-Zmp1 (Mtb-FL-Zmp1) and derived Mtb-Zp-NT and Mtb-Zp-CT peptides were purified to homogeneity (Supplementary Figure S1-C) and used in an optimized antigen-first ELISA platform, with 100 ng coating antigen and 1:200 serum dilution providing maximal signal-to-noise ratio. The assay significantly distinguished active TB from TB-recovered and exposed contacts, supporting its diagnostic specificity. Violin plots comparing full-length Zmp1 with the two epitope-derived peptides (Mtb-Zp-NT and Mtb-Zp-CT) demonstrated clear separation between active TB and all control groups (Supplementary Figure S2). Both AUC and Cohen’s *d* confirmed superior discriminatory performance of the peptides, with large effect sizes of d≥2 for Mtb-Zp-NT and d = 1.8-2.8 for Mtb-Zp-CT, compared with d=1.3-1.6 for full-length Zmp1 across the control groups (Supplementary Table S2). These findings provided preliminary evidence of disease-specific serological responses and justified expansion to a larger, clinically diverse cohort for Stage-2 evaluation.

### 3.3 Stage-2 *assessment: Mycobacterium tuberculosis* Zmp1-derived peptides, Mtb-Zp-CT and Mtb-Zp-NT, differentiate active pulmonary TB patients and active extra-pulmonary TB patients from control groups

Stage-2 extended the study to better represent the clinical heterogeneity of TB by distinguishing between pulmonary PTB and EPTB manifestations and incorporating non-specific infections as an additional control arm. A final study cohort of 875 participants: 457 with active TB and 418 controls was arrived at. Based on clinical and microbiological criteria, participants were stratified into six groups: PTB (N=347), EPTB (N=110), TBR (N=90), NSI (N=104), HHC (N=128), and HC (N=96).

Among PTB cases, antibody titers to all three antigens were significantly higher than in any control group (Wilcoxon p<0.001 for all comparisons). Violin plots demonstrated clear separation between PTB and controls, with Mtb-Zp-CT showing the greatest discrimination and minimal overlap across groups, indicating stronger immunogenicity and superior diagnostic resolution (Figure 2).

**Fig 2.**
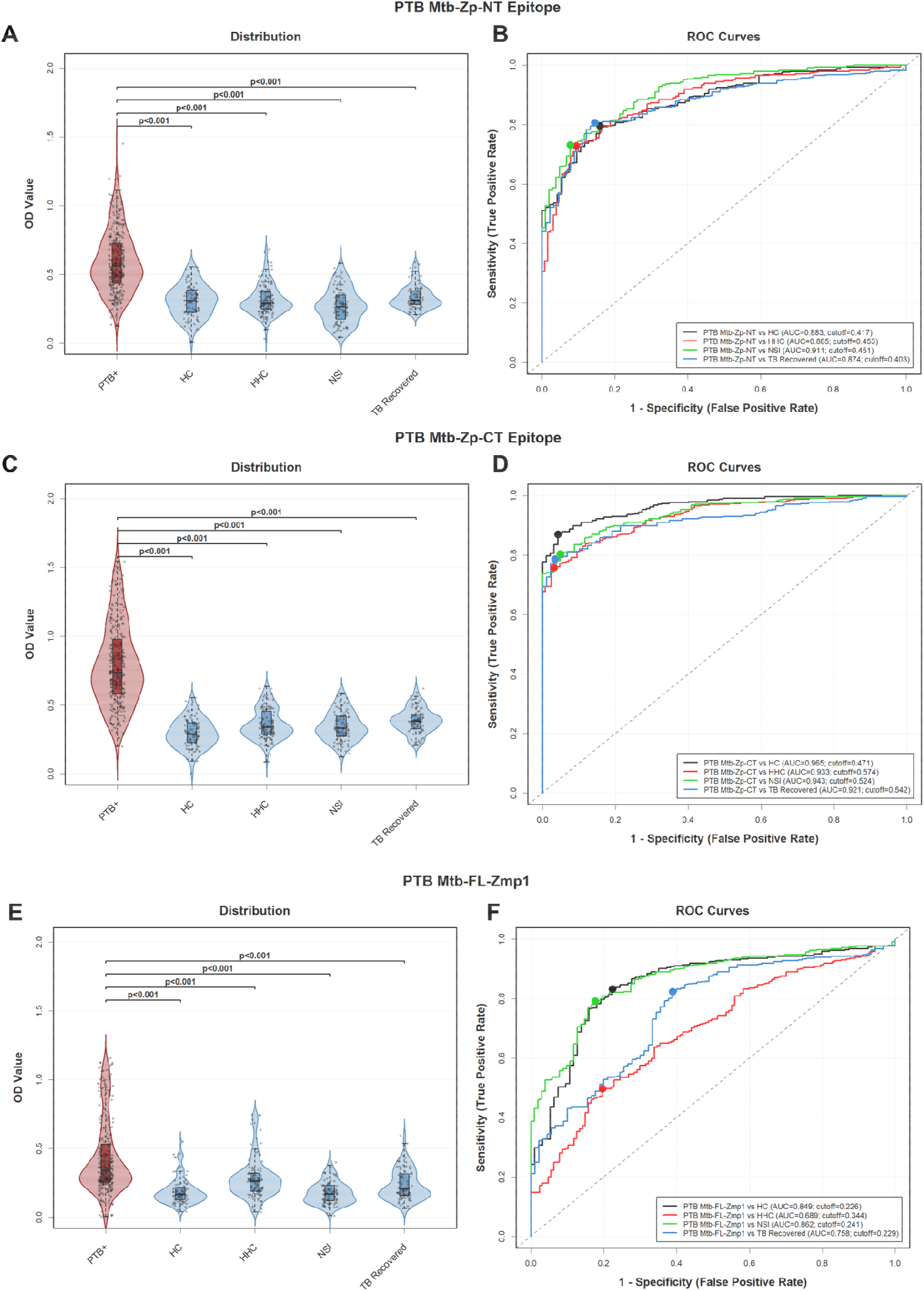
Comparative diagnostic performance of Zmp1-derived peptides and the full-length antigen in pulmonary tuberculosis. (A, C, E) Violin plots showing the distribution of ELISA-based absorbance (450 nm) values for Mtb□Zp□NT, Mtb□Zp□CT, and full□length Zmp1 protein, respectively, in sera from pulmonary TB (PTB) patients compared with healthy controls (HC), household contacts (HHC), non□specific infections (NSI), and TB□recovered individuals. Each diluted (1:200) serum sample was tested in triplicate, and the p-value was calculated by the Wilcoxon rank□sum test (**p** < 0.001). (B, D, F) Receiver operating characteristic (ROC) curves evaluating the diagnostic accuracy of each antigen. **Abbreviations in Fig 2** **Mtb-Zp-NT**: *Mycobacterium tuberculosis* Zinc Metallo Protease-1 N-terminal; **Mtb-Zp-CT**: *Mycobacterium tuberculosis* Zmp-1: Zinc Metallo Protease-1 C-terminal; **Mtb-Zp-FL**: *Mycobacterium tuberculosis* Zinc Metallo Protease-1 Full Length protein; **TB**: Tuberculosis; **N**: Sample number; **NAAT**: Nucleic Acid Amplification Test; **ELISA**: Enzyme-Linked Immuno Sorbent Assay; **HC**: Healthy Control; **HHC**: House Hold Contacts; NSI: non-specific infection; **ATT**: Anti-Tuberculosis Therapy; **PTB**: Pulmonary Tuberculosis; **ROC**: Receiver Operating Characteristics; **AUC**: Area Under Curve

Receiver-operating-characteristic (ROC) analyses confirmed these findings. Both epitope-based antigens showed consistently high diagnostic accuracy (AUC>0.8), whereas full-length Zmp1 displayed more variable performance (AUC=0.69-0.87) (Figure 2F). Mtb-Zp-CT achieved the highest AUC values across all PTB comparisons (0.92-0.97) (Figure 2D), outperforming Mtb-Zp-NT (0.87-0.91) (Figure 2B) and full-length Zmp1 (0.69-0.85). Optimal cut-offs derived using the Youden index maximised sensitivity and specificity for each antigen. The Youden index identified optimal diagnostic cutoffs for each antigen (Supplementary Table S3, Figures 2 and 3). For PTB (Figure 2), Mtb-Zp-CT cutoffs were 0.47-0.57 across control groups, with a sensitivity of 76-87% with specificity above 95% across control groups. Mtb-Zp-NT performed well at lower thresholds (0.40-0.45) with balanced metrics of sensitivity of 73-81% and specificity of 84-92%. Collectively, the integration of violin plot distributions, non-parametric statistical testing, and ROC-based metrics demonstrates that Zmp1-derived peptides, particularly Mtb-Zp-CT, provide robust serological discrimination of PTB from diverse control groups with enhanced sensitivity and specificity. These findings identify Mtb-Zp-CT as a strong candidate biomarker for the development of improved serological and point-of-care diagnostic platforms for PTB.

**Fig 3.**
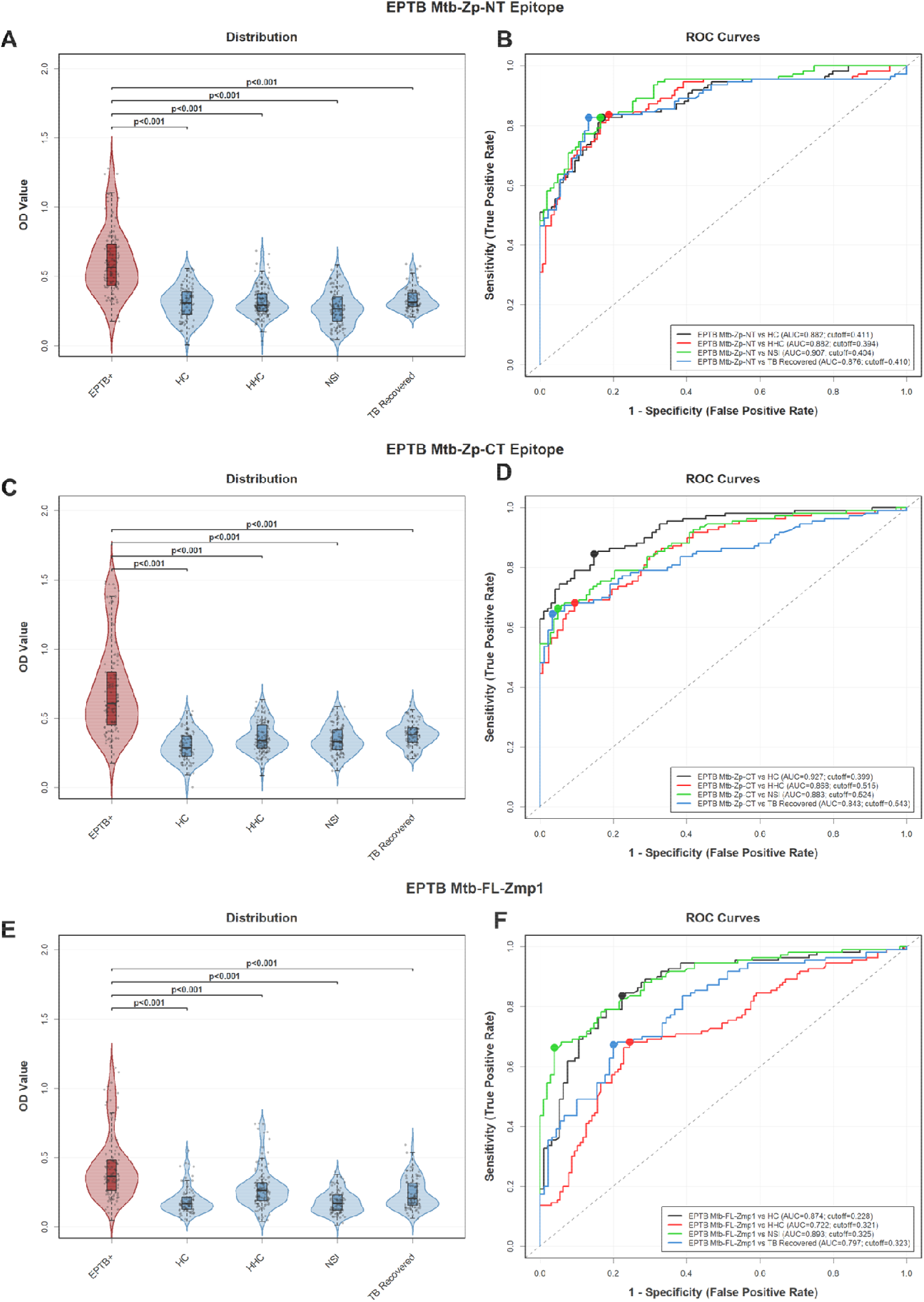
Comparative diagnostic performance of Zmp1-derived peptides and the full-length antigen in extra-pulmonary tuberculosis. (A, C, E) Violin plots depicting ELISA-based absorbance (450 nm) distributions for the Mtb□Zp□NT, Mtb□Zp□CT, and full□length Zmp1 protein, respectively, in sera from extra□pulmonary TB (EPTB) patients compared with household contacts (HHC), healthy controls (HC), non□specific infections (NSI), and TB□recovered individuals. Each diluted (1:200) serum sample was tested in tripl cate, and the p-value was calculated by the Wilcoxon rank□sum test (**p** < 0.001). (B, D, F) ROC curve analysis demonstrating the diagnostic accuracy of each antigen for EPTB. **Abbreviations in Fig 3** **Mtb-Zp-NT**: *Mycobacterium tuberculosis* Zinc Metallo Protease-1 N-terminal; **Mtb-Zp-CT**: *Mycobacterium tuberculosis* Zmp-1: Zinc Metallo Protease-1 C-terminal; **Mtb-Zp-FL**: *Mycobacterium tuberculosis* Zinc Metallo Protease-1 Full Length protein; **TB**: Tuberculosis; **N**: Sample number; **NAAT**: Nucleic Acid Amplification Test; **ELISA**: Enzyme-Linked Immuno Sorbent Assay; **HC**: Healthy Control; **HHC**: House Hold Contacts; NSI: non-specific infection; **ATT**: Anti-Tuberculosis Therapy; **EPTB**: Extra-Pulmonary Tuberculosis; **ROC**: Receiver Operating Characteristics; **AUC**: Area Under Curve

Given the strong performance of the Zmp1-derived peptides in PTB, we next assessed their diagnostic utility in EPTB using peripheral blood, aiming to establish a non-invasive assay for a TB disease form in which conventional diagnostics frequently fail. Using the same ELISA platform, we evaluated sera from clinically confirmed EPTB patients and compared responses with the four control groups: HHC, HC, NSI and TBR. Antibody titres to all three antigens, Mtb-Zp-CT, Mtb-Zp-NT, and full-length Zmp1, were significantly higher in EPTB than in any control group (Wilcoxon p<0.001) (Figure 3). Violin plots demonstrated clear separation across all comparisons, with Mtb-Zp-CT (Figure 3C) with higher distinction and minimal overlap with controls, indicating better immunoreactivity. ROC analyses further supported the diagnostic value of the peptides (Supplementary Table S3). Mtb-Zp-CT achieved the highest performance against healthy controls (AUC=0.93) (Figure 3D) and maintained robust accuracy across all other control groups (AUC>0.84). Similarly, Mtb-Zp-NT showed strong discrimination (AUC ∼0.89) (Figure 3B), whereas full-length Zmp1 displayed more variable performance (AUC=0.72-0.89) (Figure 3F). These findings indicate that both peptide-based antigens reliably distinguish EPTB from diverse control populations. In EPTB (Figure 3), Mtb-Zp-CT showed high specificity (85-97%) and sensitivity of 65-85% at cutoffs of 0.40-0.54, while Mtb-Zp-NT showed a consistent performance across all comparisons with a cutoff of 0.39-0.41, sensitivity 83-84% and specificity 81-87%.

Together, these results demonstrate that the Zmp1-derived peptides, especially Mtb-Zp-CT, provide strong serological discrimination of both PTB and EPTB using peripheral blood, supporting their potential as non-invasive biomarkers.

### 3.4 DeLong test and Bootstrap analyses for the comparative assessment of Mtb-Zp-CT and Mtb-Zp-NT on the same cohort

The DeLong test demonstrated significant differences in diagnostic performance between antigens (Supplementary Table S5). In PTB, Mtb-Zp-CT consistently outperformed both full-length Zmp1 (all p<0.001) and Mtb-Zp-NT (p=0.014 to p<0.001; Supplementary Figure S3). Mtb-Zp-NT performed better than full-length Zmp1 for household contacts (p<0.001) and individuals with non-specific infections (p=0.041), but not for healthy controls (p=0.222), indicating comparable performance in this subgroup.

In EPTB (Supplementary Figure S4, Supplementary Table S5), no significant differences were observed between the two peptides for healthy controls (p=0.079), non-specific infections (p=0.767), or TB-recovered patients (p=0.26), suggesting equivalent diagnostic performance. Both peptides, however, significantly outperformed full-length Zmp1 in household contacts (p<0.001), supporting the value of peptide-based antigens in distinguishing active disease from exposure-related responses.

Bootstrap validation confirmed the stability of these findings (Supplementary Figures S5–S10, Supplementary Table S4). In EPTB, Mtb-Zp-NT and Mtb-Zp-CT showed robust generalisability, with test AUCs of 0.89-0.91 and 0.85-0.93, respectively. In PTB, Mtb-Zp-CT again demonstrated the higher performance (AUC=0.92-0.97; accuracy >81%), whereas Mtb-Zp-NT achieved AUCs of 0.87-0.91.

Pooling all controls into a single negative group further sharpened discrimination. For PTB, Mtb-Zp-CT achieved an AUC=0.94 (cut-off 0.52; sensitivity 79%; specificity 94%; accuracy 86%), outperforming Mtb-Zp-NT (AUC=0.89; accuracy 81%) and full-length Zmp1 (AUC=0.78; accuracy 71%). In EPTB, both peptides performed better (Mtb-Zp-CT AUC=0.88; Mtb-Zp-NT AUC=0.89), where Mtb-Zp-NT showed higher sensitivity (82% vs 64%) and Mtb-Zp-CT yielded higher specificity (93% vs 83%). Diagnostic performance remained consistent whether controls were analyzed individually (Figure 4 A-F) or pooled (Figure 4G).

**Fig 4.**
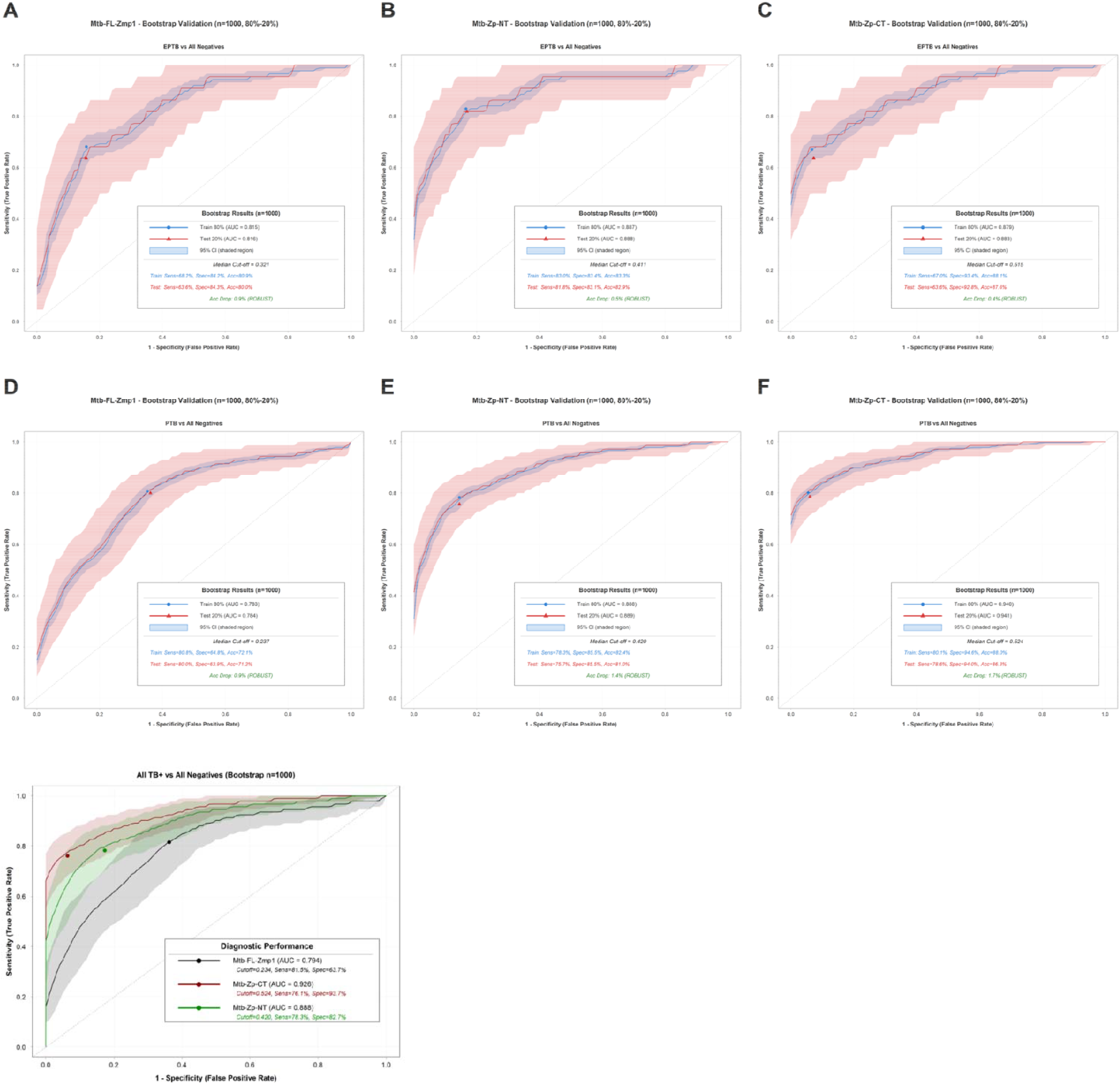
Bootstrap validation of Zmp1-derived epitopes and full-length antigen for serological discrimination of pulmonary and extra-pulmonary tuberculosis. Receiver operating characteristic (ROC) curves from bootstrap validation (N=1000 iterations, 80%-20% train-test split) show the diagnostic performance of three antigens in distinguishing TB patients from all negative controls (healthy controls, household contacts, non-specific infections, and TB-recovered individuals). Top row (A-C): ROC curves for extra-pulmonary tuberculosis (EPTB) detection using (A) Mtb-FL-Zmp1 with AUC = 0.815-0.816, (B) Mtb-Zp-NT with AUC = 0.887-0.888, and (C) Mtb-Zp-CT with AUC = 0.879-0.883. Bottom row (D-F): ROC curves for pulmonary tuberculosis (PTB) detection using (D) Mtb-FL-Zmp1 (AUC = 0.783 - 0.784), (E) Mtb-Zp-NT (AUC = 0.888-0.889), and (F) Mtb-Zp-CT (AUC = 0.940 - 0.941). Blue lines represent training set performance, red lines indicate test set performance, and shaded regions denote 95% confidence intervals. (G) distinguishing all tuberculosis-positive cases (PTB and EPTB combined) from all negative controls, including healthy controls, household contacts, non-specific infections, and TB-recovered individuals. Solid lines represent median ROC curves from test sets, with shaded regions indicating 95% confidence intervals across bootstrap iterations; the diagonal dashed line denotes random classification. **Abbreviations in Fig 4** **Mtb-Zp-NT**: *Mycobacterium tuberculosis* Zinc Metallo Protease-1 N-terminal; **Mtb-Zp-CT**: *Mycobacterium tuberculosis* Zmp-1: Zinc Metallo Protease-1 C-terminal; **Mtb-Zp-FL**: *Mycobacterium tuberculosis* Zinc Metallo Protease-1 Full Length protein; **TB**: Tuberculosis; **N**: Sample number; **NAAT**: Nucleic Acid Amplification Test; **ELISA**: Enzyme-Linked Immuno Sorbent Assay; **HC**: Healthy Control; **HHC**: House Hold Contacts; NSI: non-specific infection; **ATT**: Anti-Tuberculosis Therapy; **PTB**: Pulmonary Tuberculosis; **EPTB**: Extra-Pulmonary Tuberculosis

Bootstrap analysis of the combined TB-positive cohort (Supplementary Table S4) confirmed the superior performance of Mtb-Zp-CT, which achieved a median test AUC of 0.93 (cut-off 0.524; sensitivity 76.1%; specificity 93.7%), significantly outperforming full-length Zmp1 (AUC=0.79) and Mtb-Zp-NT (AUC=0.89). Mtb-Zp-NT showed balanced discrimination (sensitivity 78.3%; specificity 82.7%), whereas full-length Zmp1 demonstrated high sensitivity (81.5%) but poor specificity (63.7%).

Taken together, these analyses confirm that the Zmp1-derived peptides, particularly Mtb-Zp-CT, provide superior, stable serodiagnostic performance across diverse clinical scenarios. Their ability to distinguish both PTB and EPTB from high-risk groups, including household contacts and individuals who have recovered from TB, underscores their potential as refined biomarkers for point-of-care diagnostics.

## 4 Discussion

Effective humoral immunity depends on coordinated B-cell and CD4□ T-cell interactions, particularly when cognate epitopes are co-localised on the same antigen [25]. This “linked recognition”promotes antigen uptake, MHC II presentation, and focused T-cell help [26,27], collectively driving affinity maturation and sustained antibody responses. These principles increasingly inform rational antigen design for both vaccines and diagnostics. In TB, this is especially pertinent, as earlier commercial serological assays showed poor diagnostic performance, attributed largely to the use of full-length or poorly characterised antigens susceptible to cross-reactivity and inconsistent epitope presentation [28]. Reflecting these limitations, the World Health Organisation does not endorse serology for the diagnosis of PTB or EPTB, underscoring the continued need for accurate, evidence-based diagnostic tools. Nevertheless, harnessing TB-antigen-specific host immune responses holds promise for enabling early and accurate TB detection, offering a viable screening strategy in resource-limited settings and contributing to effective TB control globally.

As we know, despite advances in molecular diagnostics, TB, particularly EPTB, remains challenging to diagnose in resource-limited settings due to sputum unavailability, impractical invasive sampling, and inadequate laboratory infrastructure. Unlike pulmonary TB, EPTB lacks a defined anatomical focus, making specimen selection inherently difficult and complicating evidence-based diagnosis. The host immune response, readily accessible through peripheral blood, offers a practical alternative. A rapid, non-invasive serological assay would therefore address this diagnostic gap by enabling timely detection and facilitating early treatment initiation. We explored this possibility using Zmp1 (Rv0198c), a virulence factor [29,30], which we had identified in our previous study as an Mtb antigen that elicited strong humoral responses in PTB and EPTB patients, including smear-negative disease [31], supporting its relevance as a marker of active infection.

Our epitope mapping strategy was guided by the principle that robust antibody responses depend on cognate B-cell and CD4□ T-helper cell cooperation. Following antigen recognition by the B-cell receptor, antigen is internalized, processed, and presented on MHC class II molecules to CD4□ T cells, which provide help via CD40–CD40L interactions and cytokines (e.g., IL-4, IL-21, IFN-γ), driving B-cell activation, class switching, affinity maturation, and memory formation [12,32]. Accordingly, epitopes that combine or spatially link B-cell determinants with CD4□ T-helper epitopes are more likely to elicit strong, durable antibody responses. This rationale underpins the selection of overlapping epitope regions for improved immunogenic and diagnostic potential, as isolated B-cell epitopes often generate weaker and transient responses in the absence of T-cell help. A multi-layered immunoinformatics pipeline identified two immunodominant Zmp1 regions enriched for linear and conformational B-cell epitopes with predicted high-affinity MHC II binding. The C-terminal domain exhibited the highest epitope density, consistent with evidence that domain-focused constructs outperform full-length proteins in serodiagnostic applications [33,34]. When tested for their biomarker potency in a clinically heterogeneous cohort of 875 individuals, both Zmp1-derived peptides showed strong diagnostic performance, with Mtb-Zp-CT consistently outperforming Mtb-Zp-NT and full-length protein. High AUCs in pulmonary TB, together with minimal reactivity in HHC, TBR and NSI, suggest that Zmp1-specific antibodies reflect active disease rather than latent infection or prior exposure. Notably, Mtb-Zp-CT also effectively discriminated EPTB, a diagnostically challenging form characterized by low bacillary burden and reliance on invasive sampling.

These findings contrast sharply with the poor performance of earlier commercial serological tests, which lacked biologically informed antigen selection and were subsequently withdrawn by WHO [10]. Our results support renewed interest in rational antigen discovery and demonstrate that epitope-refined peptides can achieve high specificity even in settings with overlapping infectious syndromes. Compared with urine LAM, the only WHO-endorsed point-of-care test, but one largely restricted to advanced HIV infection [35], a serum-based Zmp1 assay offers broader applicability across HIV-negative individuals and EPTB. The statistical framework used here further strengthens confidence in the translational potential of these biomarkers. ROC analysis, Youden-derived thresholds, and extensive bootstrap sampling confirmed the stability and generalisability of diagnostic performance across subgroups. Mtb-Zp-CT consistently showed the highest accuracy, particularly when control groups were pooled to reflect real-world diagnostic settings. The strong antibody titers against our peptides give confidence in the ELISA sensitivity to be recapitulated in a lateral flow assay (LFA), with the expectation that the antibody titers in 10-20 ul of plasma will be sufficient for detection. This is an ongoing effort in our lab, with promising preliminary results supporting its translation into LFA.

In summary, the immunodominant peptide identified in this study emerges as a strong candidate biomarker aligned with WHO priorities for accessible, accurate TB diagnostics. Its ability to detect active disease, including EPTB, with high specificity and sensitivity positions it as a promising target for next-generation point-of-care assays.

## Data Availability

All data generated and analysed during this study are included in this published article [and its supplementary information files]. Requests for further information and resources should be directed to the corresponding authors, Prof. Sharmistha Banerjee and Dr Pramod Rajaram Somvanshi.

## Statements and Declarations

### Ethics approval and consent to participate

All study procedures were conducted in accordance with the ethical standards of the respective institutional research committees. Ethical approval for the study was obtained from the Institutional Ethics Committees of the University of Hyderabad (UH/IEC/2022/287), MNR (EC/NEW/INST/2023/TE/0316/1425) and BMMRC (1057/BMMRC/2021/IEC approval). Written informed consent was obtained from all participants before sample collection, and all data were anonymised to ensure confidentiality.

### Consent for publication

All authors confirm that they have reviewed the final version of the manuscript and provided their full consent for its publication.

### Competing interests

The authors declare no competing interests. A patent has been filed for the peptide as a biomarker described in the manuscript.

### Funding

We thank the Indian Council of Medical Research for funding to SB and VLV (5/8/5/21/ITRC/Diag/2021/ECD-1).

### Role of the funding source

The funders had no role in study design, data collection, data analysis, data interpretation, or writing of the manuscript.

### Authors’ contributions

OSZ: Conceptualization, experimentation, sample processing, data analyses, first draft, review and editing SY,: Conceptualization, Experimentation, Immunoinformatics based peptide prediction, first draft and review; KC: Biostatistical Analyses, first draft and review; AVG: experimentation, sample processing; RP: experimentation; MA: Clinical investigation of patients; NVSK, DKP, VVRE: samples collection & documentation, first draft and review; CC: sample collection, documentation and review; DS: Lateral flow assay design; KM: first draft, analysis, reviewing, editing, proofing; VLV: Study cohort design, supervising sample collection and documentation, proofreading; PRS: Data analyses and statistics, first draft, review and editing; and SB: Conceptualization, supervising, study design, data analyses, review, editing and fund sourcing.

## Acknowledgments

Omkar Shriram Zade thanks CSMNRF (Maharashtra) for JRF and SRF. Sriram Yandrapally thanks IoE-PDF. SB thanks DST BRICS grant (DST/ICD/BRICS/PilotCall2/MRP-TB (G)), DST-FIST to the Department of Biochemistry (SR/FST/LS-II/2023/1172), the DBT-BUILDER grant to the School of Life Sciences (BT/INF/22/SP41176/2020), and the Institution of Eminence-supported projects RC1-20-017 and RC4-21-012 to Sharmistha Banerjee and IoE to the University of Hyderabad MHRD (F11/9/2019-U3(A)) for infrastructure support to the Department of Biochemistry, School of Life Sciences, and University common facilities are acknowledged. We acknowledge the use of AI tools (Grammarly, ChatGPT, and Co-Pilot) for language editing, grammar improvement, and polishing. We acknowledge technical help from Abhishek Guggila, Jaya Bharati and the supporting staff of Mahavir and MNR hospitals. We acknowledge the clinical support by the Chief Medical Officer, UoH, Health Centre.

## Supplemental information

Supplementary Methods

Figures S1-10

Supplementary Tables 1-5

## Notes

### Competing Interest Statement

The authors have declared no competing interest.

### Author Declarations

The Institutional Ethics Committee of the University of Hyderabad, the Institutional Ethics Committee of MNR Medical College, and the Institutional Ethics Committee of BMMRC each gave ethical approval for this work. Written informed consent was obtained from all participants prior to sample collection, and all data were anonymised to maintain confidentiality.

